# Chronic Pain in Canadian Children and Adolescents: A National Population-Based Analysis

**DOI:** 10.64898/2026.08.17.26360594

**Authors:** Justine Dol, Christine T. Chambers, Jennifer A. Parker, Brittany Cormier, Kathryn A. Birnie

## Abstract

**Background:** Chronic pain affects approximately 20% of children and youth worldwide and is associated with mental and physical health impacts. Canada-specific data on the prevalence of chronic pain in children and youth are limited, highlighting the need for current high-quality population-based estimates

**Aims:** The aim of this study is to provide national estimates of self-reported chronic pain among Canadian children and youth by pain type (headache stomach ache, backache), sex (female, male), age group (5-11, 12-17 years) and province or territory.

**Methods:** Publicly available data were used from the 2019 Canadian Health Survey on Children and Youth (CHSCY), a population-based survey conducted by Statistics Canada using a nationally representative sample of Canadian children and youth

**Results:** Overall, headaches were the most commonly reported pain type (15.4%), followed by stomach aches (12.5%), and backaches (11.1%). Prevalence was consistently higher among females than males and among youth than children, with youth girls reporting the highest prevalence across all pain types. Prevalence also varied geographically, with some of the highest estimates observed in the Atlantic Provinces.

**Conclusions:** Chronic pain affects substantial proportions of Canadian children and youth with disparities observed by pain type, sex, age, and geography. These findings under score pediatric chronic pain as an important public health issue and highlight the need for equity-oriented approaches that address the needs of populations experiencing the greatest burden.

## Background

Chronic pain is experienced by approximately 20.8% of children and youth worldwide (Chambers et al., 2024). Chronic pain is defined by the International Association of the Study of Pain (ISAP) as “pain in one or more anatomic regions that persists or recurs for longer than 3 months and is associated with significant emotional distress or significant functional disability (interference with activities of daily life and participation in social roles) and that cannot be better explained by another chronic pain condition” (Nicholas et al., 2019, p. 1004). Chronic pain is also recognized in the International Classification of Disease (ICD)-11, defined as “an unpleasant sensory and emotional experience associated with, or resembling that associated with, actual or potential tissue damage. Chronic pain is pain that persists or recurs for longer than 3 months” (MG30) (World Health Organization, 2022). Pain is multifaceted and can present as headaches, abdominal pain, back pain, and musculoskeletal pain and may occur independently of injuries or underlying medical conditions (Nicholas et al., 2019).

When left unmanaged, chronic pain in youth is linked to poorer mental health outcomes (Dudeney et al., 2024; Noel et al., 2016; Wrona et al., 2021), increased school absenteeism (Norton & Southon, 2020; Sato et al., 2007), greater socioeconomic disparities (Murray et al., 2020), higher substance use (Pielech et al., 2020; Ripon & Maleki, 2025), and reduced physical activity and higher obesity rates (Kichline et al., 2019; Wilson et al., 2010). Approximately 50% of youth with chronic pain continue to experience chronic pain in adulthood (Feinstein et al., 2025; Horst et al., 2014; Sillanpää & Saarinen, 2018). Prevalence rates of chronic pain vary by pain type, with one in four young people reporting chronic headaches and one in five experiencing chronic back or abdominal pain (Chambers et al., 2024). Differences have also been found based by age, with the prevalence of chronic pain increasing across childhood and adolescence, and by sex, with chronic pain disproportionately affecting girls compared to boys (Chambers et al., 2024; Nilles et al., 2023).

Although there is a recent meta-analysis examining worldwide prevalence data on chronic pain in children and adolescents (Chambers et al., 2024), there is a dearth of data describing its prevalence in the Canadian context. Among the 448 data points available between 2009 and 2023 included in that meta-analysis, only 7 (1.6%) were from Canada. These data came from two multi-national studies, both of which used the Health Behaviour in School-aged Children (HBSC): WHO Collaborative Cross-National survey dataset (Chambers et al., 2024; Gobina et al., 2019; Swain et al., 2014). Gobina et al. found that multi-site pain was the most common pain experienced in Canadian adolescents (20.5%), followed by headaches (11.4%), backaches (8.1%), stomach aches (4.0%). Swain et al. found much higher rates – 61% of Canadian children reporting headaches, 58.1% experienced stomach aches, and 45.8% experienced backaches. Previous data published from the National Longitudinal Survey of Children and Youth found that up to 30% of Canadian youth experience frequent headaches, while up to one in five reported frequent stomach aches and backaches (Stanford et al., 2008). A more recent population-based study using the 2019 Canadian Health Survey on Children and Youth (CHSCY) estimates the prevalence of frequent headaches among youth to be 6.1% (Nilles et al., 2023). However, this study examined only headaches and did not consider other types of chronic pain, such as stomach aches and back aches. Taken together, these findings highlight the limited and fragmented evidence base on pediatric chronic pain in Canada. Therefore, further research is needed to provide contemporary national estimates of chronic pain by type, age, sex, and geography.

### Aim

The aim of this study was to provide national estimates of self-reported chronic pain among Canadian children and youth by pain type (headache, stomach ache, backache), sex (female, male), age group (5-11, 12-17 years) and geography (province and territory).

### Objectives

1. Estimate the prevalence of headaches, stomach aches, and backaches among Canadian children and youth using a population-based sample,
2. Estimate the prevalence of headaches, stomach aches, and backaches by sex (female and male), age group (5-11 years and 12-17 years), and geography (province and territory).

## Methods

### Context

Canada operates a publicly funded, universal health care system established under the Canada Health Act, which sets national principles of comprehensiveness, universality, and accessibility but delegates the organization, funding, and delivery of care the provincial and territorial health systems (Canada Health Act, 1985). As a result, the structure and availability of specialized services, including pediatric chronic pain care, are determined jurisdictionally and vary considerably across the country (T. Palermo, 2025). Health Canada’s “An Action Plan for Pain in Canada” identifies children and youth as disproportionately affected by pain and calls for improved, systematic data collection to monitor pain incidence, prevalence, impact, and outcomes over time (Canadian Pain Task Force, 2021), indicating that information on chronic pain in children and youth is a national priority. By producing national estimates disaggregated by pain type, sex, age, and geography, this study establishes the baseline surveillance data needed to inform policy, target service planning, and monitor progress over time in addressing chronic pain among Canadian children and youth.

### Study Design and Data Source

This descriptive study used publicly available data from the 2019 Canadian Health Survey on Children and Youth (CHSCY) (Government of Canada, 2019). The CHSCY is a national, population-based survey conducted by Statistics Canada in collaboration with Health Canada, the Public Health Agency of Canada, provincial and territorial ministries of health and other governmental and academic partners. It provides comprehensive, cross-sectional data on the physical and mental health of Canadian children and youth and was designed to be representative of the Canadian population aged 1–17 years as of January 31, 2019. The Canada Child Benefit database was used to construct the survey sampling frame. The CHSCY survey collects data on areas that impact the physical and mental health of children and youth living in Canada (e.g., chronic conditions, physical activity, nutrition, recreational screen time, sleep) as well as demographic variables (e.g., sex, age group, geography). The main objectives of the CHSCY are to collect detailed health data on children and youth within and across Canada, inform the development and evaluation of health policies and programs, and support research on child and youth health. To date, the CHSCY has been administered at two time points: 2019 and 2023.

### Data

Anonymized, aggregate partial data from the 2019 survey relevant to the aim of this study were obtained from Statistics Canada Table 13-10-0763-01, “Health characteristics of children and youth aged 1 to 17 years 2019.”, released on July 23, 2020. 2023 data was publicly not available at time of data analysis. It includes data from 15 health indicators and three demographic variables. Of interest to this study were three health indicators related to types of chronic pain (headaches, stomach aches, backaches) and three demographic variables (sex, age group, geography). These sociodemographic characteristics as available through the Statistics Canada report are able to provide a national descriptive epidemiology of pediatric chronic pain, recognizing that other sociodemographic variables may be relevant but were not available in the publicly available data tables. Ethics review was not required for this study as all data used in this analysis were publicly available.

Geographic information includes separate data for each of the 10 Canadian provinces and combined data for the three territories. Age group data included children aged 1 to 4 years and youth aged 5 to 17 years, further stratified into 5-11- and 12-17 years. Data on chronic pain were available only for participants aged 5-17 years. Data by sex were limited to two categories, male and female. No data on gender was available.

The three chronic pain indicators in the 2019 CHSCY survey were defined as children and youth aged 5–17 years who experienced headaches, stomach aches, or backaches once per week or more during the previous six months, with responses reported by the person most knowledgeable for children aged 5–11 years (e.g., caregiver) and self-reported by youth aged 12–17 years. This definition differs slightly from both the IASP’s and ICD-11’s definition of chronic pain (Nicholas et al., 2019; World Health Organization, 2022). Response options were binary (yes/no). Participants could report more than one pain condition, therefore, estimates for each pain type are not mutually exclusive.

### Participants

The 2019 CHSCY surveyed children and youth living in Canada using a stratified sampling design that included participants from all 10 provinces and the three territories in Canada aged of 1 to 17 years as of January 31, 2019. Excluded from the survey by Statistics Canada were children and youth living on First Nations reserves and other Indigenous settlements in the provinces, children and youth living in foster homes, and institutionalized children and youth (Government of Canada, 2019). Data were collected between February 11, 2019 and June 28, 2019 and included 92,170 respondents (raw units). Survey responses were voluntary completed either online or by telephone with a Statistics Canada interviewer. The data used in this study were collected via questionnaires administered to the person most knowledgeable about the selected child or youth. For children 5-11, this means the survey was typically completed by a caregiver or other adult (i.e., proxy reporter) and self-reported by youth aged 12–17 years. Additional details on the 2019 CHSCY survey are available elsewhere (Government of Canada, 2019).

### Statistical analysis

Descriptive statistics were used to calculate weighted prevalence estimates (%) and 95% confidence intervals for children reporting chronic pain by type (headache, stomach ache, backache) in the past 6 months, and stratified by sex (female and male), age group (5–11, 12– 17 years, and overall), and geography (provinces and territories). Survey sampling weights were provided by Statistics Canada to generate nationally representative estimates.

## Results

The prevalence estimates of chronic pain among children and youth aged 5-17 years are shown in Table 1. Headaches were reported most frequently (15.4%), followed by stomach aches (12.5%) and backaches (11.1%). Females reported a higher prevalence of all pain types than males. For headaches, 19.9% of females reported chronic pain compared with 11.2% of males. For stomach aches, 16.3% of females reported chronic pain compared with 8.9% of males. Finally, for backaches, 13.8% of females reported chronic pain compared with 8.5% of males.

**Table 1.** Percentage of Canadian Children and Youth (Aged 5–17 Years) Reporting Headaches, Stomach Aches, and Backaches During the Previous 6 Months (2019), Overall and Stratified by Age Group and Sex.

|  | **Headaches**<br>**% (95% CI)** | **Stomach aches**<br>**% (95% CI)** | **Backaches**<br>**% (95% CI)** |
| --- | --- | --- | --- |
| **Overall** |  |  |  |
| 5-17 years | 15.4 (14.9-16.0) | 12.5 (12.0-13.0) | 11.1 (10.6-11.5) |
| 5-11 years | 6.3 (5.8-6.8) | 9.4 (8.8-10.0) | 1.86 (1.6-2.1) |
| 12-17 years | 26.0 (25.0-27.0) | 16.3 (15.5-17.1) | 22.0 (21.0-23.0) |
| **Male** |  |  |  |
| 5-17 years | 11.2 (10.5-11.8) | 8.9 (8.4-9.5) | 8.5 (7.9-9.1) |
| 5-11 years | 5.2 (4.6-5.9) | 7.4 (6.7-8.1) | 1.7 (1.4-2.1) |
| 12-17 years | 18.2 (17.1-19.5) | 10.8 (9.9-11.8) | 16.6 (15.4-17.8) |
| **Female** |  |  |  |
| 5-17 years | 19.9 (19.1-21.0) | 16.3 (15.5-17.1) | 13.8 (13.2-14.5) |
| 5-11 years | 7.3 (6.6-8.2) | 11.5 (10.5-12.5) | 2 (1.7-2.5) |
| 12-17 years | 33.0 (33.0-37.0) | 22.0 (21.0-23.0) | 28.0 (26.0-29.0) |

**Table 2.**
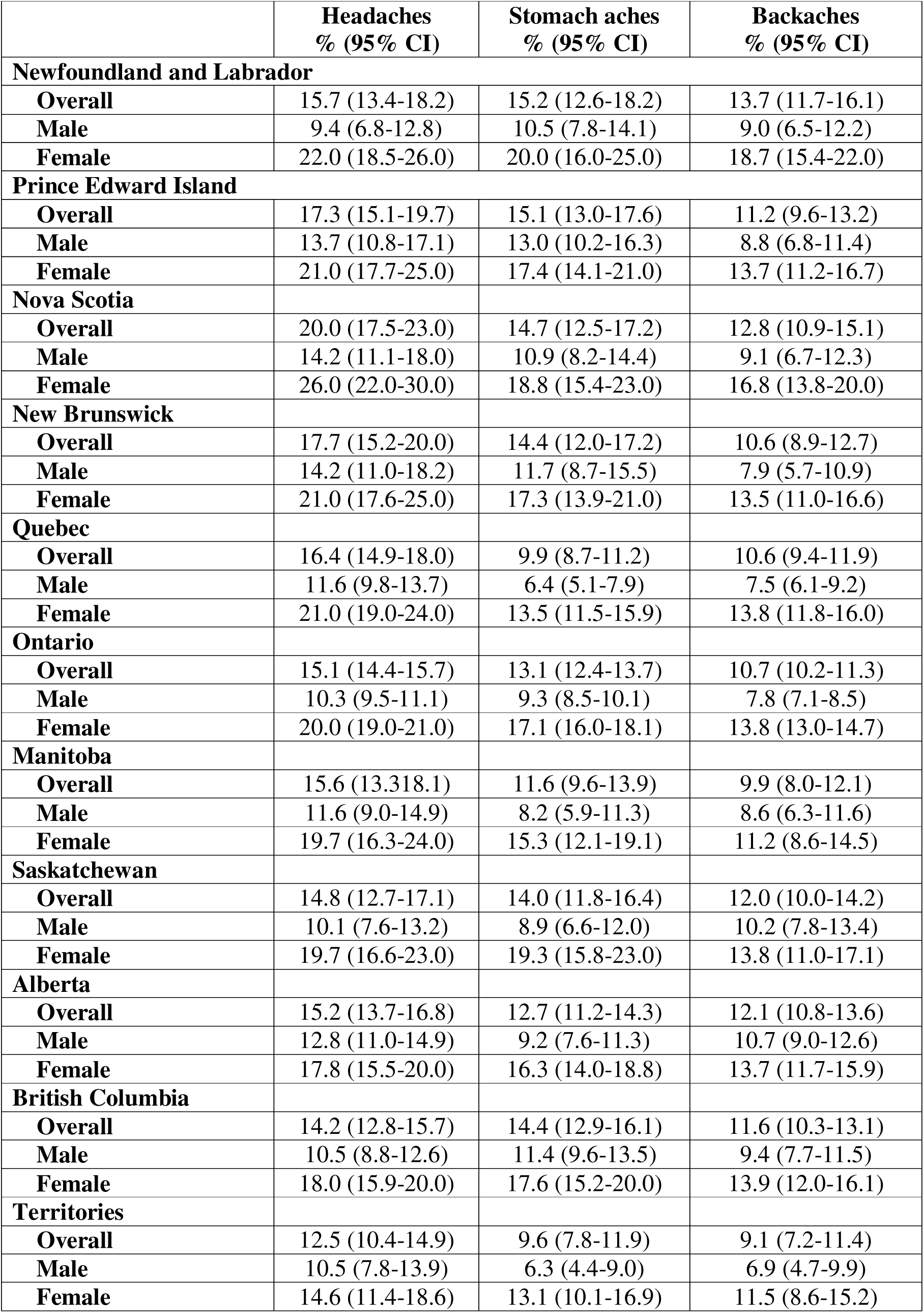
Percentage of Canadian Children and Adolescents (Aged 5–17 Years) Reporting Headaches, Stomach Aches, and Backaches During the Previous 6 Months (2019), by Province and Territory and Stratified by Sex.

When stratified by age group, youth aged 12-17 years reported a higher prevalence of headache (26.0%), backaches (22.0%), and stomach aches (16.3%) than children aged 5-11 years (Table 1). This pattern was consistent across sexes. The highest percentages of reported pain were by females aged 12-17 years for headaches (33.0%), stomach aches (22.0%), and backaches (28.0%).

By geography, Nova Scotia had the highest prevalence of headaches among children and youth aged 5-17 years followed by New Brunswick (17.7%, see Figure 1). Newfoundland and Labrador had the highest prevalence of both stomach aches (15.2%) and backaches (13.7%), followed by Prince Edward Island (15.1%) and Nova Scotia (12.8%), respectively. When further stratified by sex, females in Nova Scotia had the highest reported prevalence of headaches (26.0%), whereas females in Newfoundland and Labrador had the highest prevalence of stomach aches (20.0%) and backaches (18.7%). The lowest prevalence of headaches was reported among males in Newfoundland and Labrador (9.4%), whereas males in the territories had the lowest prevalence of both stomach aches (6.3%) and backaches (6.9%).

**Figure 1.**
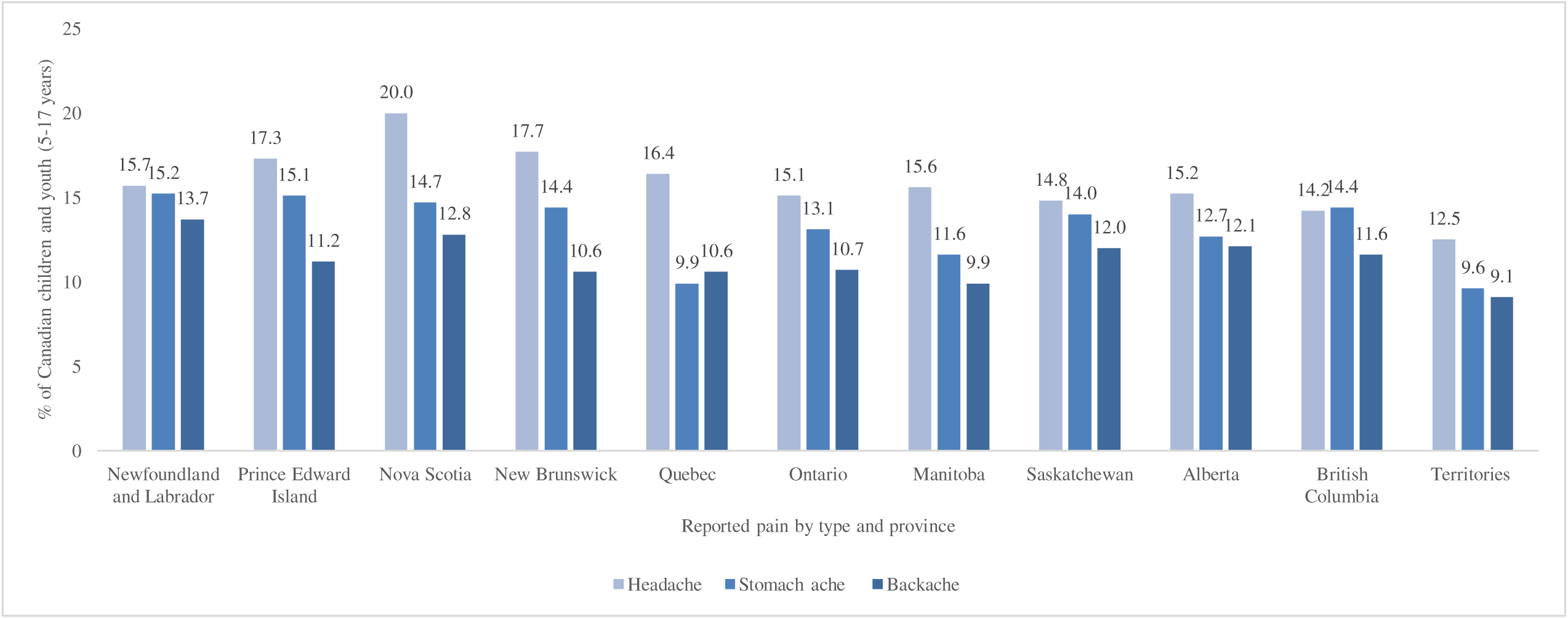
Percentage of Canadian Children and Adolescents (Aged 5–17 Years) Reporting Headaches, Stomach Aches, and Backaches During the Previous 6 Months (2019), by Province and Territory

**Figure 2.**
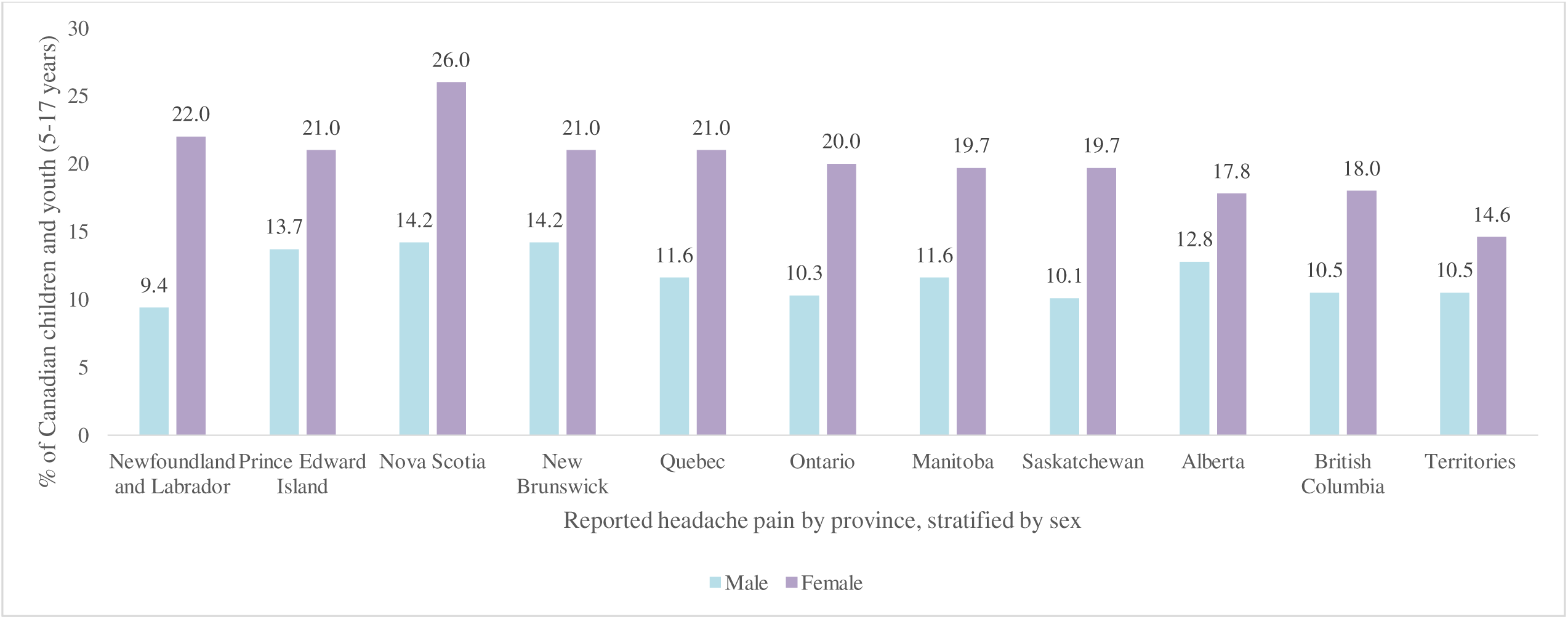
Percentage of Canadian Children and Adolescents (Aged 5–17 Years) Reporting Headaches During the Previous 6 Months (2019), by Province and Territory and Stratified by Sex

**Figure 3.**
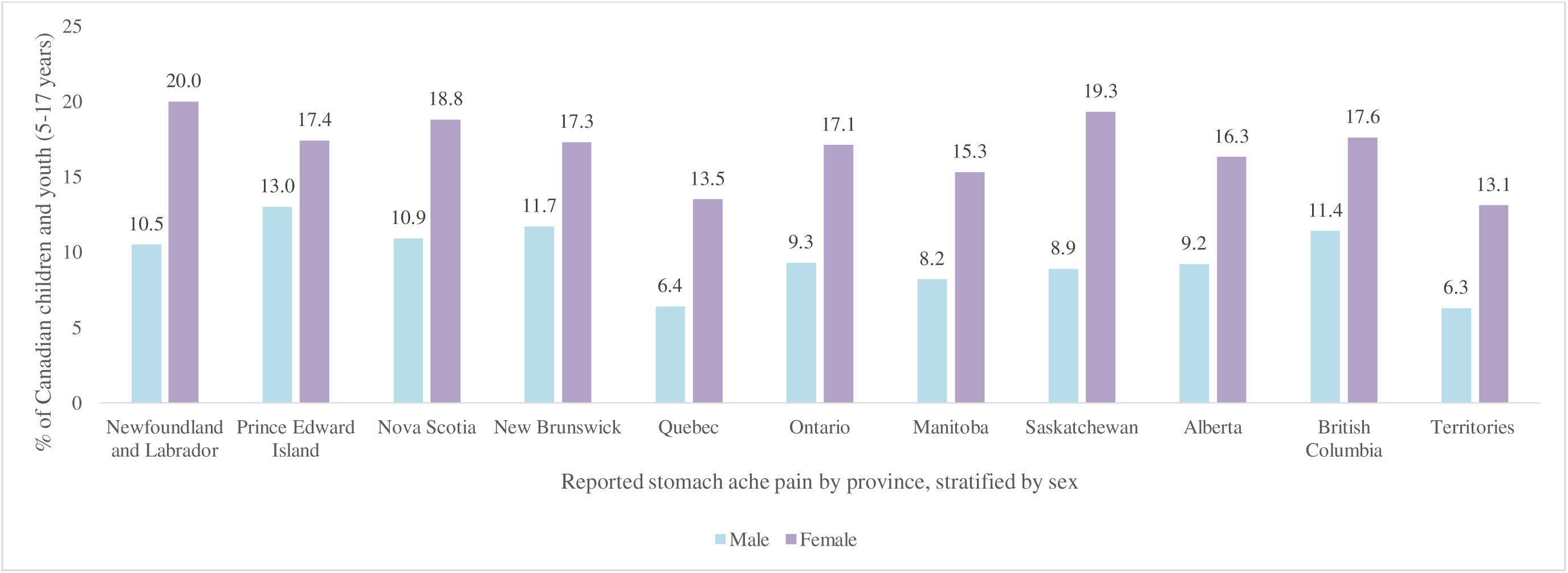
Percentage of Canadian Children and Adolescents (Aged 5–17 Years) Reporting Stomach Aches During the Previous 6 Months (2019), by Province and Territory and Stratified by Sex

**Figure 4.**
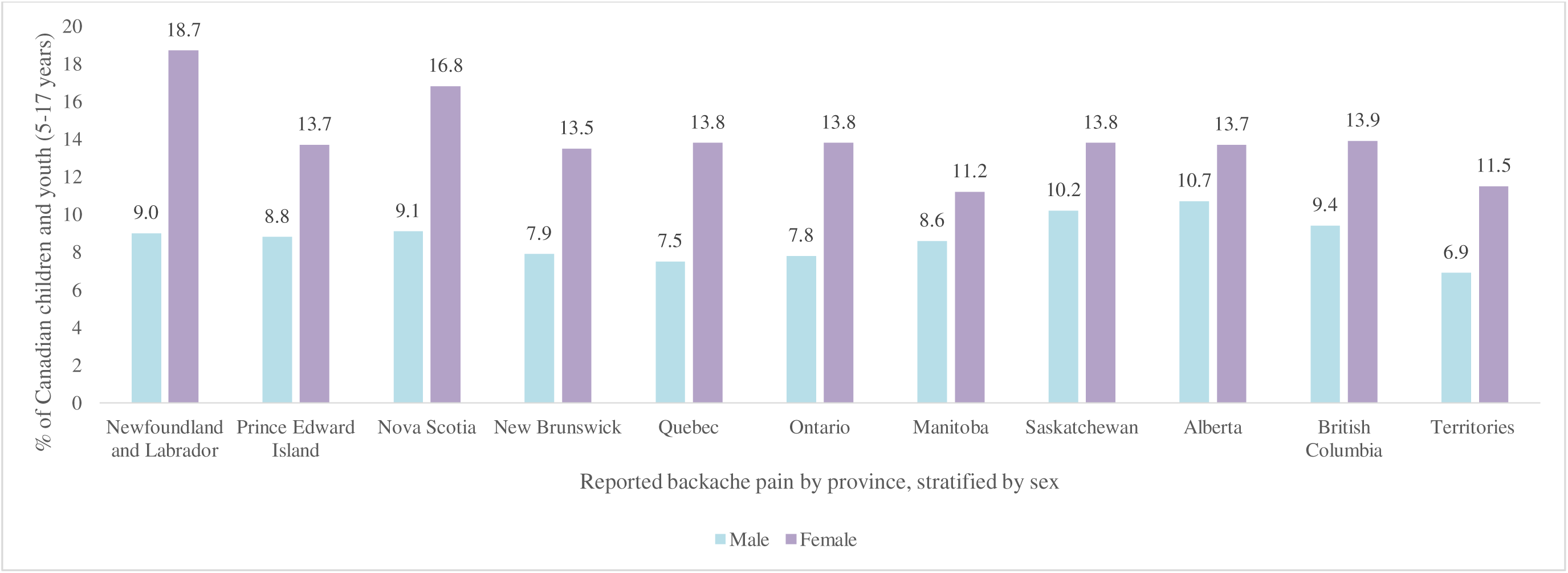
Percentage of Canadian Children and Adolescents (Aged 5–17 Years) Reporting Backaches During the Previous 6 Months (2019), by Province and Territory and Stratified by Sex

### Conclusions

This study provides a national overview of chronic pain (headaches, stomach aches, and backaches) among children and youth in Canada. While previous meta-analytic data suggests that 20.8% of children and youth experience chronic pain worldwide (Chambers et al., 2024), the estimated prevalence in Canada for these pain types ranged from 11.1% for backaches to 15.4% for headaches as reported in this population-based survey. However, when stratified by sex, females reported a higher prevalence of all pain types than males, and when stratified by age, youth reported a higher prevalence than children, consistent with previous literature (Chambers et al., 2024). A novel contribution of this study was a comparison across provinces and territories, which showed that all of the Atlantic provinces (Newfoundland and Labrador, Prince Edward Island, Nova Scotia, New Brunswick) generally had a higher prevalence of chronic pain than other regions of Canada. Collectively, these findings underscore that chronic pain is a substantial public health concern: it is common, persistent, and detrimental not only to the well-being of children and youth but also to their families and the broader healthcare system. Chronic pain affecting at least one in seven children and youth in Canada, with clear differences by sex, age, and geography. These estimates confirm that a substantial proportion of young people live with chronic pain which is consistent with the Action Plan for Pain in Canada’s recognition of children and youth as a population of concern, and reinforcing its call for a coordinated public health response (Canadian Pain Task Force, 2021). Such a response, built on collaboration among youth and families, health professionals, decision-makers, and communities and spanning prevention, equitable access to care, and sustained surveillance, is needed to address pediatric chronic pain at the population level.

As found in a recent meta-analysis by Chambers and colleagues (2024), worldwide 23.5% of females reported headaches, 15.1% reported backaches and 11.3% reported stomach aches. This is consistent with our Canadian findings where 19.9% of females reported headaches, 13.8% reported backaches and 16.3% reported stomach aches. This higher prevalence of chronic pain in females compared to males is well documented in the literature (Chambers et al., 2024; Fillingim et al., 2009; Mogil, 2012). This difference may be linked to puberty and hormonal changes, which may influence pain sensitivity and perceptions (Li et al., 2023; Maurer et al., 2016), as well as inflammatory pathways that can differ by sex (Fang et al., 2023; Manson, 2010), potentially contributing to a greater risk of chronic pain among females. Unfortunately, the publicly available data only provides biological sex and did not consider gender, despite substantial evidence that gender and gender norms can play a role in pain expression (Alabas et al., 2012; Boerner et al., 2018; Nascimento et al., 2020). Nevertheless, the continued evidence that females experience more chronic pain than males support the need for sex-responsive pain assessment and interventions in the pediatric population, with earlier screening among girls potentially warranted.

In the current study, overall data from children and youth aged 5–17 years were further stratified into two age groups – children aged 5-11 years and youth aged 12-17 years, with adolescents reporting higher prevalence of chronic pain than children. Previous studies have found that differences in pain experience, particularly during experimental pain tasks, may emerge around 12 years of age (Boerner et al., 2014). This may be due to a variety of factors including hormonal shifts (Maurer et al., 2016) and rapid neurodevelopment (Colver & Longwell, 2013), which may influence the emergence or persistence of chronic pain during this vulnerable developmental window. Importantly, youth were more likely to complete the survey themselves, whereas responses for children aged 5–11 years were provided by the person most knowledgeable (typically a parent or caregiver). This difference in respondent may partially explain the higher prevalence observed among youth, as they may have greater awareness of their pain experience than proxy respondents. Thus, age-related differences in chronic pain prevalence may reflect not only developmental changes but also differences between self- and parent-reported data, with parents often reporting lower levels of pain than children report themselves (Hanania et al., 2024; Kamper et al., 2016). These findings should therefore be interpreted in light of differences in respondent across age groups.

Children and youth in Atlantic Canada had the highest prevalence of pain across all pain types. In a similar study comparing adult pain prevalence across Canada, Newfoundland and Labrador, Saskatchewan, and New Brunswick had the highest pain prevalence among all provinces (Zajacova et al., 2022). This pattern may be related to health disparities observed in Atlantic Canada, particularly greater rurality and higher levels of socioeconomic disadvantage, both of which have been associated with higher reported pain and other adverse health outcomes (Quon & McGrath, 2015; White et al., 2011; Zajacova et al., 2022). Atlantic Canada has a higher burden of chronic disease among adults (Mortey et al., 2025; Robitaille et al., 2012), raising the possibility that some of these disparities emerge during childhood. While these findings may reflect environmental and policy differences, evidence also suggests that pain behaviours may be modelled within families, with children experiencing chronic pain more likely to have parents with chronic pain (Higgins et al., 2015; T. M. Palermo et al., 2014; Stone et al., 2018). Given the higher prevalence of chronic pain in Atlantic Canada, future research should examine how structural inequalities contribute to these regional differences and identify opportunities for systems-level interventions to improve child and youth health.

### Strengths and Limitations

This study has several notable strengths. It draws on nationally collected data from Statistics Canada, providing a large, population-based sample that is broadly representative of Canadian children and youth. This enhances the generalizability of findings and allows for meaningful comparisons across provinces and territories. The detailed provincial analysis is a particular strength, as few studies have examined pediatric chronic pain patterns at this level of geographic granularity in Canada, and is relevant given geographical disparities and relevance of rurality to accessing speciality pediatric chronic pain care (T. Palermo, 2025; Sahaym & Groenewald, 2026).

At the same time, several limitations should be considered. Because the survey was not designed specifically to measure chronic pain, its case definition differs from the internationally accepted definition put forward by the IASP, limiting comparability with other studies, including the meta-analysis conducted by Chambers et al. While the inclusion of common pain types such as headache, stomach ache, and backache aligns with prior epidemiological work, the survey does not capture the full spectrum of chronic pain conditions (e.g., disease-specific or widespread pain syndromes), potentially underestimating overall burden. Accordingly, the prevalence estimates should be interpreted in this context. The publicly available data were limited to binary sex categories and did not include gender, limiting the ability to examine gender-diverse experiences of chronic pain and underscoring the need for more inclusive measurement and reporting in future national surveillance efforts. Finally, Indigenous populations are not fully represented in the CHSCY, highlighting an important gap in understanding chronic pain inequities in Canada, as previous work has highlights that Indigenous individuals may have a higher prevalence of pain symptoms than the general population (Jimenez et al., 2011; Julien et al., 2018). Other social and structural determinants of health with emerging evidence demonstrating relevance to pediatric chronic pain prevalence were not measured, such as immigration status (Roman-Juan et al., 2025, 2026), race and racism (Hood et al., 2023), neighbourhood characteristics (Vandeleur et al., 2024), and household food insecurity (Tham et al., 2023), and are worth considering in a public health response.

### Future Research

Several important directions for future research emerge from these findings. First, further statistical modeling is needed to more fully examine the intersection of sex, age, and geography, including formal interaction testing to determine whether certain subgroups experience disproportionately elevated prevalence. More granular analyses incorporating socioeconomic status, rurality, access to care would also help clarify potential structural and contextual factors underlying observed regional differences. Future research should build on these descriptive findings by examining the sociodemographic correlates and longitudinal predictors of chronic pain, including socioeconomic disadvantage, family characteristics, and other sociopolitical and structural determinants, to better understand pathways underlying observed disparities and to guide effective public health responses.

Second, longitudinal research is essential to better understand developmental would allow examination of onset, persistence, and remission patterns, as well as identification of early predictors of chronic pain that may inform targeted prevention strategies. A life-course approach is particularly important given evidence that pain in adolescence persists into adulthood and is intergenerational (Beveridge et al., 2025; Higgins et al., 2015).

Finally, given the timing of national data collection cycles, future work should explicitly examine pre- and post-COVID-19 patterns of chronic pain given demonstrated impact of the pandemic on youth with chronic pain and their families, and associated healthcare utilization (Soltani et al., 2023). The pandemic introduced substantial disruptions to schooling, physical activity, mental health, healthcare access, and social connection (Dunton et al., 2020; Loades et al., 2020; Moynihan et al., 2021; Van Lancker & Parolin, 2020), all of which may influence chronic pain risk and maintenance. Comparative analyses across survey cycles would help determine whether prevalence patterns have shifted and whether existing disparities by sex, age, or geography have widened. Together, these directions would strengthen our understanding of chronic pain epidemiology in Canadian children and youth and support more precise, equity-informed intervention strategies.

## Conclusion

Taken together, these findings underscore that chronic pain in children and youth in Canada is not evenly distributed, but varies by sex, age, and geography in ways that have meaningful clinical and policy implications. The higher prevalence among girls and youth represents critical windows of vulnerability, likely shaped by intersecting biological, psychological, and social processes. Furthermore, the elevated burden in the Atlantic provinces highlights that chronic pain may also be influenced by broader regional, geographic, and structural contexts, including access to care, socioeconomic conditions, rurality, and health system capacity. Addressing chronic pain in children and youth therefore requires a life-course, public health, and equity-oriented approach lens that recognizes populations experiencing the greatest burden, invests in accessible and inclusive care, and prioritizes regionally tailored solutions to help reduce long-term pain trajectories and their downstream impact on population health.

## Author’s contributions statement

The content and views expressed in this article are those of the authors and do not necessarily reflect those of the Government of Canada.

## Declaration of generative AI use

The authors report generative AI was not used in their research or preparation of this manuscript.

## Data deposition

This study uses data publicly available from Statistics Canada How to cite: Statistics Canada. Table 13-10-0763-01 Health characteristics of children and youth aged 1 to 17 years, Canadian Health Survey on Children and Youth 2019

## Data Availability

All data used in this study is available online at https://doi.org/10.25318/1310076301-eng

